# Dominant and rare SARS-Cov2 variants responsible for the COVID-19 pandemic in Athens, Greece

**DOI:** 10.1101/2020.06.03.20121236

**Authors:** Spanakis Nikolaos, Kassela Katerina, Dovrolis Nikolas, Bampali Maria, Gatzidou Elisavet, Kafasi Athanasia, Froukala Elisavet, Stavropoulou Anastasia, Lilakos Konstantinos, Veletza Stavroula, Tsiodras Sotirios, Tsakris Athanasios, Karakasiliotis Ioannis

## Abstract

SARS-CoV-2 (severe acute respiratory syndrome coronavirus 2) is a novel Coronavirus responsible for the Coronavirus Disease-2019 (COVID-19) pandemic. Since the beginning of the pandemic, the virus has spread in almost the entire world. Tracing and tracking virus international and local transmission has been an enormous challenge. Chains of infections starting from various countries worldwide seeded the outbreak of COVID-19 in Athens, capital city of Greece. Full-genome analysis of isolates from Athens’ Hospitals and other healthcare providers revealed the variety of SARS-CoV-2 that initiated the pandemic before lock-down and passenger flight restrictions. The present work may serve as reference for resolving future lines of infection in the area and Europe especially after resumption of passenger flight connections to Athens and Greece during summer of 2020.

## Introduction

SARS-CoV-2 (severe acute respiratory syndrome coronavirus 2) is a novel Coronavirus responsible for the Coronavirus Disease-2019 (COVID-19) pandemic [1]. SARS-CoV-2 was first reported in December 2019, when a pneumonia outbreak occurred in Wuhan City, Hubei Province, China [2]. The infection rapidly spread throughout mainland China and subsequently to all continents, in at least 212 countries. On March 11^th^ -2020, the World Health Organization (WHO) declared COVID-19 a pandemic [3]. So far, there are over 6.2 million confirmed SARS-CoV-2 cases worldwide that have caused more than 373,000 deaths. The clinical manifestations of COVID-19 vary notably among affected individuals. Most patients develop mild to moderate flu-like symptoms, however some cases progress to severe pneumonia and acute respiratory distress syndrome (ARDS) that can be fatal [4]. The association of many of the initial cases with the Huanan Seafood Wholesale Market in Hubei, a wildlife animal market, suggested a zoonotic origin of the virus [5].

Coronaviruses (CoVs) belong to the *Coronavirinae* subfamily of *Coronaviridae* family and *Nidovirales* order. Alpha and betacoronaviruses infect only mammals, including humans, whereas gamma, and delta coronaviruses infect mainly birds, although some of them can also harm mammals [6]. In humans, CoVs cause primarily respiratory and enteric infections that can be either mild, such as the common cold, or more severe manifestations such as bronchitis and pneumonia [7]. SARS-CoV-2 is the seventh characterized coronavirus that infect humans. Already known coronaviruses CoV-HKU1, CoV-NL63, CoV-OC43 and CoV-229E induce mostly mild diseases [7] while SARS-CoV (severe acute respiratory syndrome coronavirus) and MERS-CoV (Middle East respiratory syndrome coronavirus) are highly pathogenic and caused large outbreaks of severe respiratory disease during 2002–2003 [8], and in 2012 and onwards [9], respectively.

SARS-CoV-2 is a large, enveloped virus, belonging to the genus *Betacoronavirus*. The viral genome is a ~30 kb single-stranded, positive-sense RNA, encoding approximately 9860 amino acids. Two open reading frames (ORFs), ORF1a and ORF1b, located at the 5’ terminus of the genomic RNA encode 16 non-structural proteins (nsps), involved in virus replication and possibly in the evasion of host immunity. In addition, the viral genome encodes four structural proteins (spike protein [S], envelope protein [E], membrane protein [M], and nucleocapsid protein [N]), and several accessory proteins [10, 11]. The envelope spike glycoprotein mediates receptor binding on the host cell and plays an essential role in determining host tropism [12]. SARS-CoV-2 uses the ACE2 (angiotensin-converting enzyme 2) receptor to enter human cells, as does SARS-CoV [13].

Molecular characterization of the SARS-Cov2 pandemic begun from the very first isolate to arise in Hubei, China [2] and has already yielded more than 10000 complete genome sequences worldwide [14]. SARS-Cov2, although not rapidly evolving (possibly due to the ability of its nsp14 exonuclease [15] to correct replication errors), has already shown divergence from the initial isolate that has differentially spread around the globe. Various methodologies have divided, locally or internationally, the virus isolates into two or more clades/lineages [14, 16-18]. Tracing virus variability through full genome sequencing apart from its use as an analytical tool to assist the epidemiological analysis of the pandemic may also provide an approach to tracking antigenic diversity that will assist towards the assessment of potential antigenic drift of the virus during outbreaks as well as towards vaccine development.

Greece is one of the countries in Europe least affected by the current pandemic, mainly due to its effective response strategy that included tracing and isolating cases and contacts and effective institution of social distancing measures. The first imported case was reported on February 26, 2020, while gradual lockdown measures were imposed between the 10^th^ and 23^rd^ of March 2020 [19]. The greater area of the capital city of Athens, the most densely populated area in Greece, was the main epicentre of the pandemic encompassing the majority of cases and considerable spread of the virus in the community. Our study focused on the genomic characterization of a number of isolates from a number of healthcare facilities in the Metropolitan area of Athens that included 14 hospitals and other diagnostic or health care providers. The aim of the study was to analyse the entire viral genomes through next generation sequencing and to analyse heterogeneity and dominant variants circulating and spreading throughout the capital during the first month of the outbreak. These variants form a representative group of viruses that seeded Athens before and during gradual lockdown. The analysis focused on links between countries with high viral prevalence and Greece and possible lines of infection among positive cases in the country.

## Materials and Methods

### Samples and viruses

Between March 5 and April 4 2020, oropharyngeal and nasopharyngeal swabs were submitted for molecular diagnosis for SARS-Cov2 virus to the Laboratory of Microbiology, Medical School, National and Kapodistrian University of Athens, designated as one of the two reference laboratories for COVID-19 in Athens, Greece. During the aforementioned period 421 positive samples were acquired, representing approximately 25% of country’s cases until April 4 2020. A list of 94 samples was randomly formed and curated for duplicates and family acquired infections in order to better assess viral variability (**Suppl. Table 1**).

### Extraction of RNA and Reverse Transcription real-time PCR

Nasopharyngeal or Oropharyngeal dacron swabs were collected from suspected cases with or without symptoms of Covid-19 with a history of travelling or close contact with a confirmed case. All swabs were rehydrated in 500μl PBS. RNA was isolated from 250μl of PBS rehydrated swabs in a final elution volume of 70μl, using the automated Promega’s© Maxwell Viral Total Nucleic Acid Purification Kit. For the detection of SARS-CoV-2 RNA, Genesig’s© COVID-19 CE-IVD RT Real Time PCR kit was utilized and performed according to manufacturer’s instructions starting from 8μl of eluted RNA.

### Next Generation Sequencing

Libraries were prepared using the Ion AmpliSeq Library Kit Plus according to the manufacturer’s instruction using Ion AmpliSeq SARS-CoV-2 RNA custom primers panel (ID: 05280253, ThermoFisher Scientific). In brief, the RNA library preparation involved reverse transcription using SuperScript VILO cDNA Synthesis Kit (ThermoFisher Scientific), 15-19 cycles of PCR amplification, adapter ligation, library purification using Agencourt AMPure XP (Beckman Coulter), and library quantification using Qubit Fluorometer high-sensitivity kit. Ion 530 and 540 Chips were prepared using Ion Chef and NGS reactions were run on an Ion GeneStudio S5, ion torrent sequencer (ThermoFisher Scientific).

### Bioinformatics

Quality control of Ampliseq reads, as well as, their alignment to the severe acute respiratory syndrome coronavirus 2 isolate Wuhan-Hu-1, complete genome [20], was performed within the Torrent Server of Ion Torrent S5 sequencer using default settings. The aligned reads were utilized for both reference-guided assembly and variant calling. Assembly was performed using the Iterative Refinement Meta-Assembler (IRMA) v.0.6.1 [21], that produced a consensus sequence for each sample using a >50% cut-off for calling single nucleotide polymorphisms. IRMA utilizes multiple steps of alignment, variant calling and consensus building by capitalizing on multiple allele frequency confidence intervals and read depth. Aligned reads were validated through the Integrative Genomics Viewer (IGV) v.2.5.3 [22]. Even though this approach was very accurate for single nucleotide substitutions of high allele frequency we wanted to explore the variants in our reads in their entirety. For this reason we used the LoFreq v.2.1.4 software [23] to identify even low frequency variants present in our samples. LoFreq is very sensitive and can detect variants that exist even in a few aligned reads while evaluating those based on quality metrics (cut-off *p*= *0.01)*. Taking into consideration that coronavirus genome contains long stretches of homopolymers, quasispecies analysis was focused on single nucleotide polymorphisms. Variants were annotated using SnpEff v.4.5covid19 [24] in order to be assigned to a specific viral protein. SnpEff utilizes a genomic feature file (gff) that contains all the information on the viral protein structure and intervals on the reference sequence. Positions of synonymous or non-synonymous (missense) variants were plotted on viral genome.

The phylogenetic tree (cladogram) encompassing the isolates of the present study was constructed using FastTree v.2.1.10 [25] and visualized in R using the ggtree package [26]. The phylogenetic tree encompassing European and European-related international isolates was constructed using FastTree v.2.1.10 [25] as implemented for NextStrain platform [14]. Tree calculations were based on maximum-likelihood method with branch size taking into consideration temporal data [25]. Country annotation of isolate clusters took into consideration the dominant (100%) country confidence rate of the main nodes where the strains of the present study are located.

Lineage assignment was achieved via the Pangolin COVID-19 Lineage Assigner interface [18] (https://pangolin.cog-uk.io/) and custom scripts in R and python were used to handle big data and create visualizations.

## Results

The current study focused on the genomic characterization of SARS-Cov2 variants that initiated Coronavirus outbreak in Athens, Greece in 2020. It has been hypothesized that the 2020 Coronavirus epidemic in Greece was initiated via imported cases from various countries where SARS-Cov2 already circulated after its international spread from China during January and February 2020. Full genome analysis, through next generation sequencing, was performed on 94 isolates from Athens, Greece. Ninety-four samples yielded high quality complete genome sequences with coverage ranging between 99.3 and 100 %, and fold coverage ranging between 288x and 46377x (median = 3942x) (**Suppl. Figure 1**). Complete genome sequences were deposited on GISAID and NCBI databases (Genbank Accesion Numbers: MT459832-MT459925).

### Multiple lineages seeded the COVID-19 epidemic in Athens

Amongst the systems that currently divide SARS-Cov2 isolates into distinct clades/lineages the dynamic nomenclature system presented by Rambaut et al. serves as a reliable international platform focusing on the most widespread variants and their descendants [18]. Lineage analysis of the 94 full-length genomes resulted in the assignment of each isolate to one of the currently circulating lineages (**Figure 1**). Isolates fell into 9 out of 51 lineages (May 2020). The dominant lineage was B.1 followed by lineages B.2.1 and B.2 that presented much lesser representation (**Figure 1**). A map of nucleotide polymorphisms for each strain highlighted common and differentiating genetic traces of the isolates (**Figure 1**).

**Figure 1.**
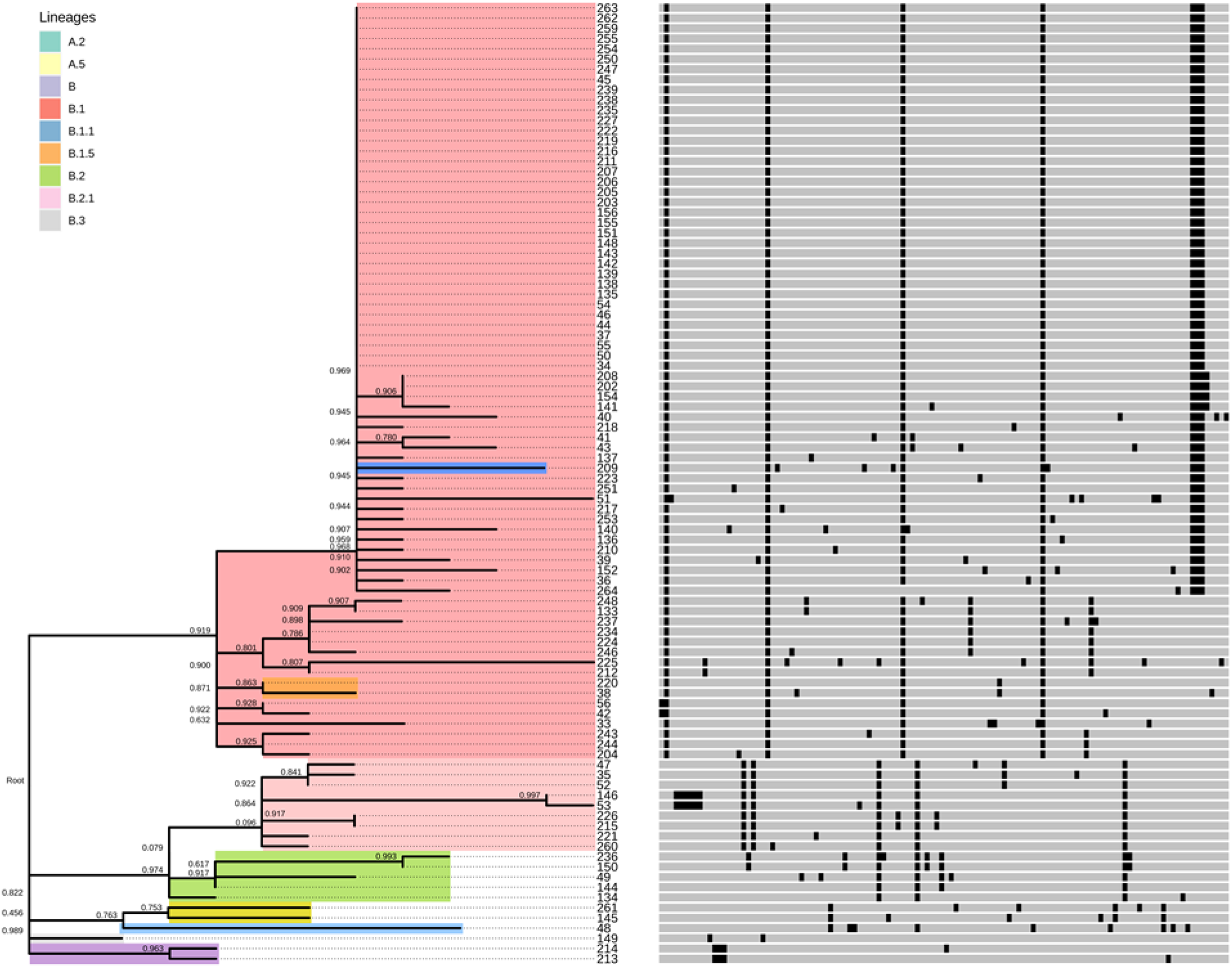
Phylogram with polymorphism profile of SARS-Cov2 variants from Athens, Greece. Tree calculations were based on maximum-likelihood method. The ratio of replicate trees in which the associated taxa clustered together in the bootstrap test (1000 replicates) are shown next to the branches. Colours represent SARS-Cov2 lineages.

### Importation of distinct variants from countries heavily affected by the pandemic

A phylogenetic tree based on maximum-likelihood analysis of European and European-related international isolates was adapted from NextStrain platform using temporal branch constrains. Designation of a country of origin on each main tree node, according to maximum (100%) country confidence rate, revealed potential importation of multiple virus variant from a variety of heavily affected countries such as the UK, Belgium and Spain (**Figure 2**). Moreover, multiple branches related to the above countries encompassed one or more virus variants, implying multiple events of virus transmission (**Figure 2**). A list of isolates from NextStrain platform that was assembled using the minimum pairwise distances (<10^-12^) between the pairs of tips that included isolates from the present study revealed a similar pattern of transmission relationships among isolates with the same genetic traits (**Suppl. Table** 2, **Suppl. Figure 3**).

**Figure 2.**
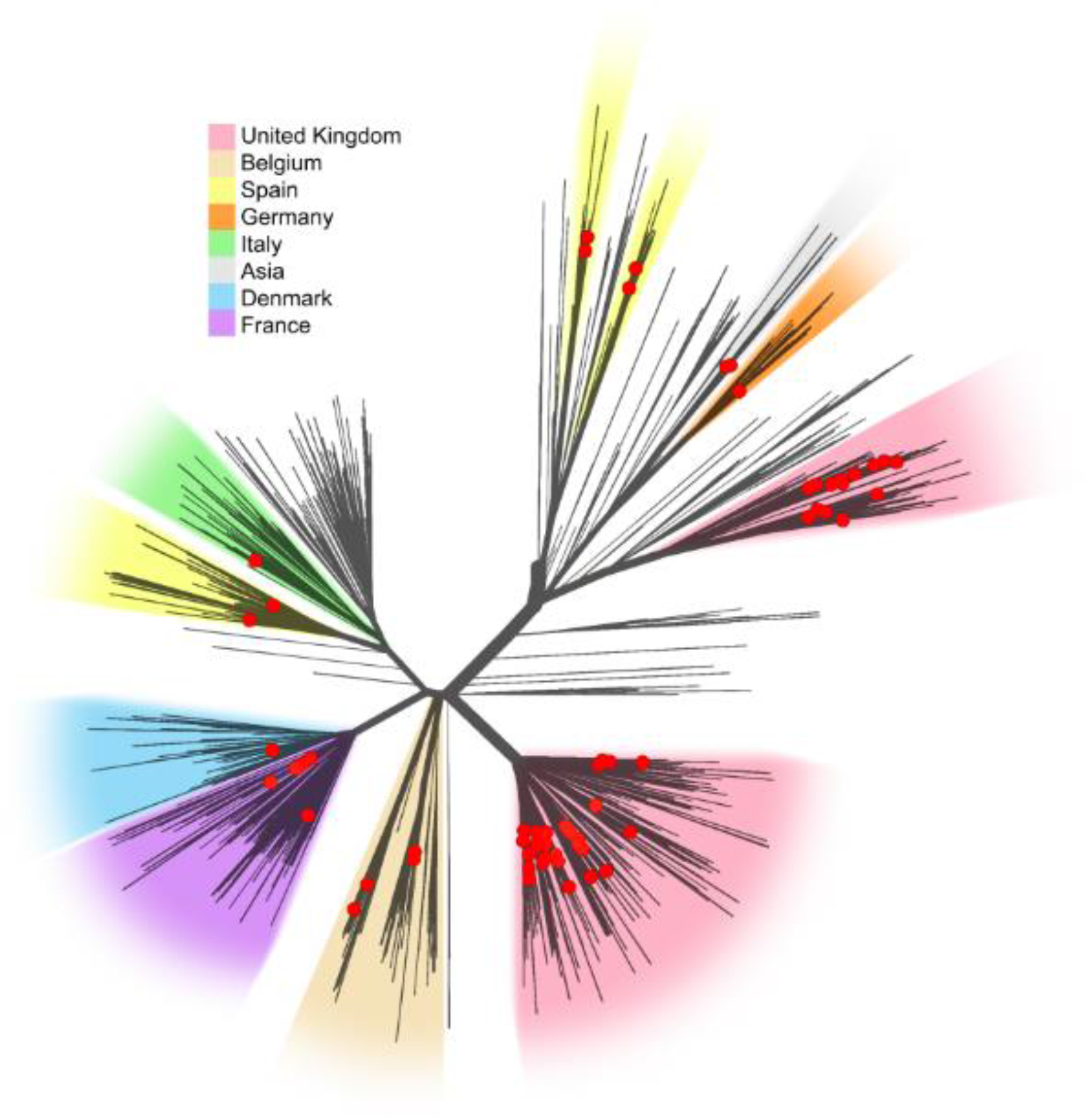
Unrooted phylogenetic tree based on temporal data of European isolates and their global links. Tree was adapted by **Next strain** (4668 complete genomes, 18/05/2020). Colours represent clusters with (100%) country confidence rate for the respective country of predicted origin.

### Non-extensive nosocomial transmission during the first month of the epidemic in Athens

Grouping isolate lineages according to the healthcare provider where virus sampling took place did not provide evidence of specific variant outbreaks in Athens’ hospitals (**Figure 3**). Distribution of B.1 lineage was even probably due to its dominant spread across Athens (**Figure 3, Suppl. Figure 4**). However, two probable hospital-acquired infections were pinpointed within B2.1 lineage. Isolate 146 was collected 16 days after the collection date of isolate 53 owing one additional mutation as compared to isolate 53. Both isolates presented unique genetic traits as compared to the rest of the isolates of B2.1 cluster as they encompassed a deletion of 6 nucleotides in the 5’ extremity of ORF1ab (**Figure 1**). Isolates 215 and 226, collected 4 days apart within a cluster of interlinked hospitals, demonstrated 100% similarity and unique genetic traits (two differentiating mutations) as compared to the rest of the isolates of B2.1 cluster. The observed similarity between the two isolated implied transmission between hospitals, possibly from a common source as the collection days are only 4 days apart (**Figure 1**).

**Figure 3.**
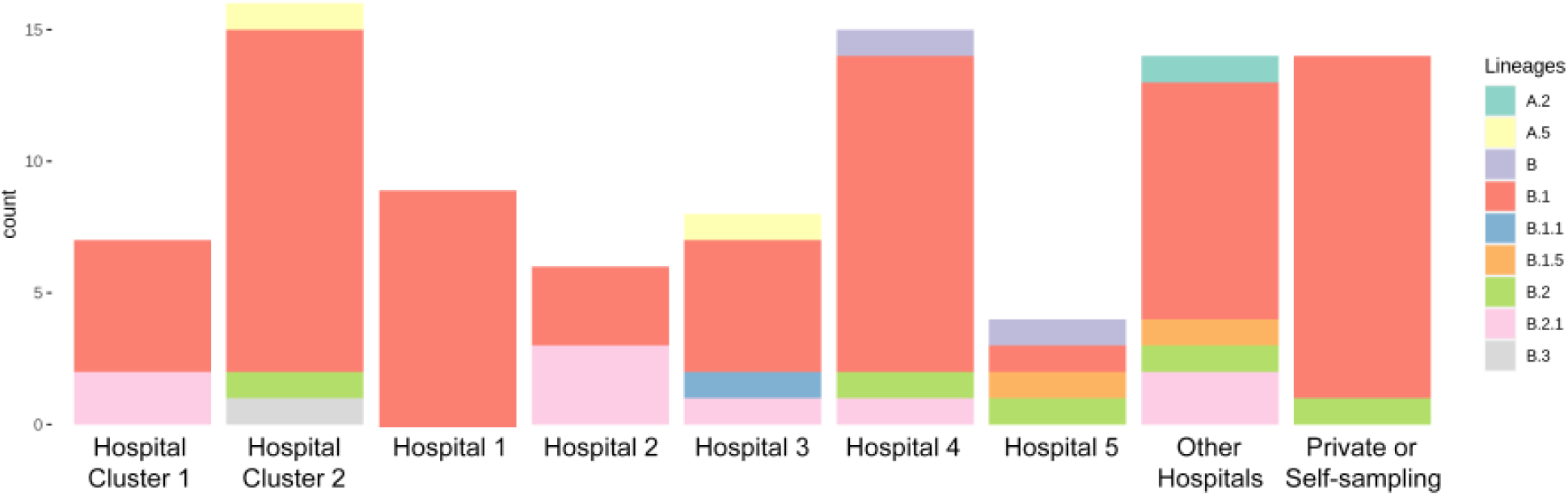
Stacked Bar Graph of lineages from samples acquired from various healthcare providers in Athens, Greece.

### Mapping of viral genetic variability on the virus genome

One hundred twenty one (121) unique dominant (>50% representation in the sample) nucleotide polymorphisms and major quasispecies (Lofreq *p= 0.01)* were inferred from the mapped reads using Lofreq method for each isolate as compared to the reference Hu-1 SARS-Cov2 strain. Nucleotide polymorphisms and major quasispecies from all the isolates of the study, represented as spikes over the viral genome, highlighted positions with high divergence potential amongst quasispecies (black spikes, **Figure 4**). Several quasispecies frequently rose in different patients signifying areas with high and low evolutionary potential. Mapping the density of unique positions of quasispecies within the 94 samples supported this notion (**Figure 4**). Quasispecies observed among Athens isolates are concentrated in hot spots located in limited areas of ORF1ab (eg. nsp3 and nsp6) and more predominantly in the downstream ORFs encoded by coronavirus subgenomic RNAs (e.g. S, ORF8 and N). High evolutionary potential regions are often hotspots for aminoacid substitutions that may have direct effect on protein functionality. Indeed amino acid substitutions presented similar clustering in the same regions of the genome (**Figure 4**).

**Figure 4.**
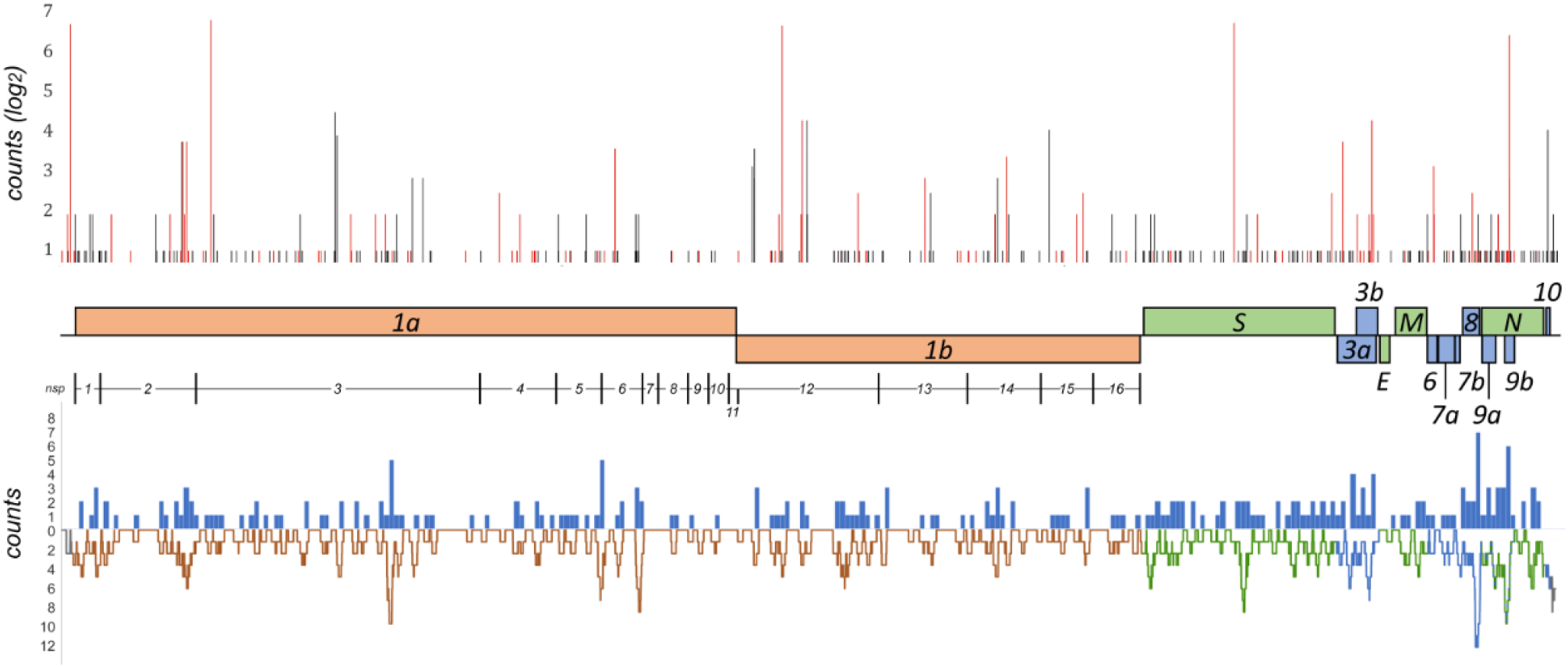
Map of SARS-Cov2 variability in Athens, Greece. Abundance of nucleotide polymorphisms (red spikes) and quasispecies (black spikes) of SARS-Cov2 variants in this report, plotted against viral genome (upper graph). Density of missense quasispecies represented by histogram of counts per 100 bases and density of all quasispecies by plotting rolling sum (frame: 100 nts). Colours represent respective regions of the viral genome (lower graph).

### Isolates with deleterious genetic traits

Five isolates presented deletions of aminoacids. Two closely related isolates (146 and 53) belonging to the B2.1 lineage presented a deletion of two amino acids in ORF1a (nsp1). This deletion has been reported previously in various isolates (e.g. MT385441, MT344956 as presented in NCBI Genbank). Similarly, two closely related isolates (213 and 214) belonging to the B lineage presented an one-amino-acid deletion in ORF1a (nsp2). This deletion has also been reported previously in various isolates (e.g. MT457399, MT451006 deposited in NCBI Genbank). An isolate (51) diverging significantly from the B.1 lineage presented a premature stop codon within ORF7a. The stop-codon-variant was the predominant variant in the sample (1179 out of 1183 reads) while the wild-type variant was only present as background (1 out of 1183 reads). Premature termination of the protein was predicted to result in the abrogation of 28 amino acids from the C terminus of ORF7a protein. Intriguingly, this moiety represents the intrinsically disordered C terminal region of SARS coronavirus ORF7a protein as implied by the 3D structure of the SARS ORF7a protein (**Suppl. Figure 5**) [27].

## Discussion

The COVID-19 pandemic first appeared in Greece on 26 February 2020 when the first COVID-19 case imported from Northern Italy, was confirmed. Although various regions in the country have been affected since, the major epicentre of the epidemic was in the greater area of the capital city of Athens. Athens accommodates almost one third of the country’s population, while it is the major economic and travelling centre of the country. Despite the fact that the first cases in various European countries appeared around the same time, Greece is one of the least affected by the pandemic countries [28]. While the epidemic is still ongoing around the globe, introduction or reintroduction of the virus in the communities could be a major threat for immunologically naïve populations especially those belonging to high risk groups. As Greece is a major summer-time destination for European and International tourists, close epidemiological surveillance of post-quarantine local SARS-Cov2 outbreaks is required in order to avoid or contain a second pandemic wave and viral reintroduction in the country.

Full genome next generation sequencing of viruses during local or widespread epidemics, (e.g. the Ebola virus epidemic), has particularly assisted as an analytical tool for the determination of conventional and unconventional transmission chains [29-31]. SARS-Cov2 pandemic is the first pandemic that has been rapidly monitored through genomic analysis of isolates around the world [14]. Phylogenetic analysis clustering described major and minor viral lineages with various degrees of prevalence from the very beginning of the pandemic. High prevalence of major lineages was assisted by major SARS-Cov2 outbreaks in Europe followed by outbreaks in the Americas [14, 28]. Following the dynamic nomenclature system by Rambaut et al., the outbreak in Athens during the first 5 weeks after the first identified case, was initiated in a greater extent by lineage B.1 viruses and at a lesser extent by lineages A2, A5, B, B1.1, B1.5, B.2, B2.1 and B.3. Worldwide dominant distribution of lineage B.1 may account for its high prevalence in Athens, while introduction of numerous other lineages implies multiple routes of virus introduction in the country [32]. Lineages did not present any clustering regarding the health care provider supporting limited hospital-acquired infections during the first 5 weeks of the epidemic in Greece. However, two cases of suspected nosocomial infections were observed, both of which are related to B2.1 lineage viruses.

Specific branches of the SARS-Cov2 phylogenetic tree in Europe encompass dominant variants from local outbreaks in European countries that were vastly affected by the pandemic. Such variants with minimal or no divergence are found amongst the Athens isolates, signifying direct transmission between countries. Athens isolates cluster in the European phylogenetic tree in multiple branches that represent genetic traits that were first observed in countries that served as transmission hubs for Europe and subsequently for the Americas [14], such as Belgium, United Kingdom, Italy and Spain. Intriguingly, Athens was mostly affected by viruses that share genetic traits with variants originating from the United Kingdom, possibly correlating with a wave of repatriation of Greeks leaving the UK during the outbreak.

Further genomic analysis of Athens SARS-Cov2 isolates revealed a rich variety (121) of dominant (>50% in a sample) nucleotide polymorphisms and multiple high and low abundance quasispecies. Quasispecies observed among Athens isolates are located in hot spots across the genome, although concentrated in the structural/accessory protein part as previously observed [14]. Genetic variations such as insertions, deletions and premature stop codons are predicted to confer significantly larger effects. In the present report 2 different amino acid deletions in ORF1a were observed both represented by two related isolates. Both deletions have been observed in isolates internationally [14]. Another strain presented a premature stop codon in ORF7a resulting in the abrogation of 28 amino acids. It is the first time that this variant is reported although others have previously reported an extensive deletion in ORF7a [33]. Such variations may represent the continuous adaptation of the virus to human cells or viral tendency towards natural attenuation [34, 35].

Next generation sequencing of full viral genomes has greatly assisted epidemiology either resolving lines of infection or assessing virus variability in the community. Rapid genetic characterization of isolates following molecular diagnosis can be a powerful analytical tool of molecular epidemiology. During the post-lockdown era of the pandemic, local outbreaks and sporadic cases should be promptly traced, especially in immunologically naïve countries like Greece. While cessation of quarantine measures is required to reduce economic effects of the pandemic, resumption of passenger flight connections to Athens and Greece may present a serious challenge in tracking sporadic cases or outbreaks. Our work delineated the pool of variants that initiated the outbreak of SARS-Cov2 in Athens and may serve as reference for resolving future lines of infection in the area and Europe.

## Data Availability

Data are publicly available is NCBI and GISAID platforms as indicated in the manuscript

https://www.gisaid.org/

https://www.ncbi.nlm.nih.gov/nucleotide/

## Acknowledgements

We would like to thank Professor Ian Goodfellow, Department of Pathology, University of Cambridge, for helpful discussions.

## Supplementary Figures

**Suppl. Figure 1.**
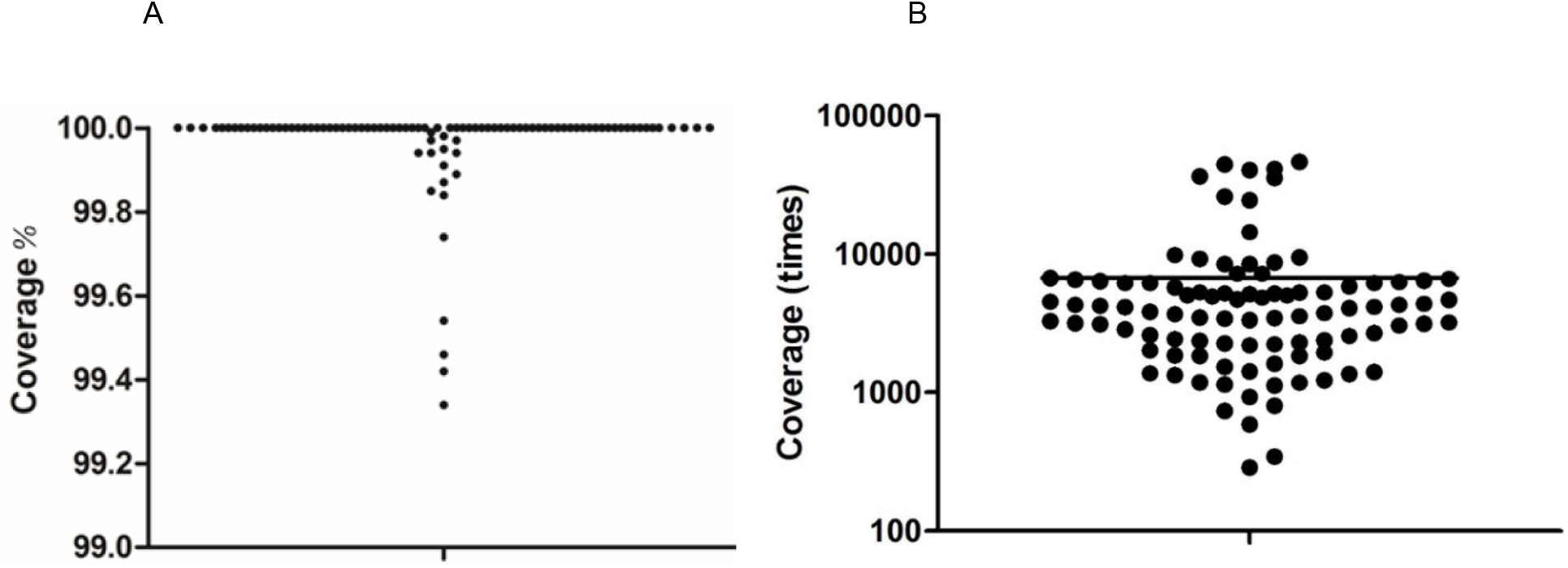
Coverage as percentage (%) of the SARS-Cov2 reference strain Hu-1 (A) and Coverage as fold (x) of the 94 complete genome sequences of the report.

**Suppl. Figure 2.**
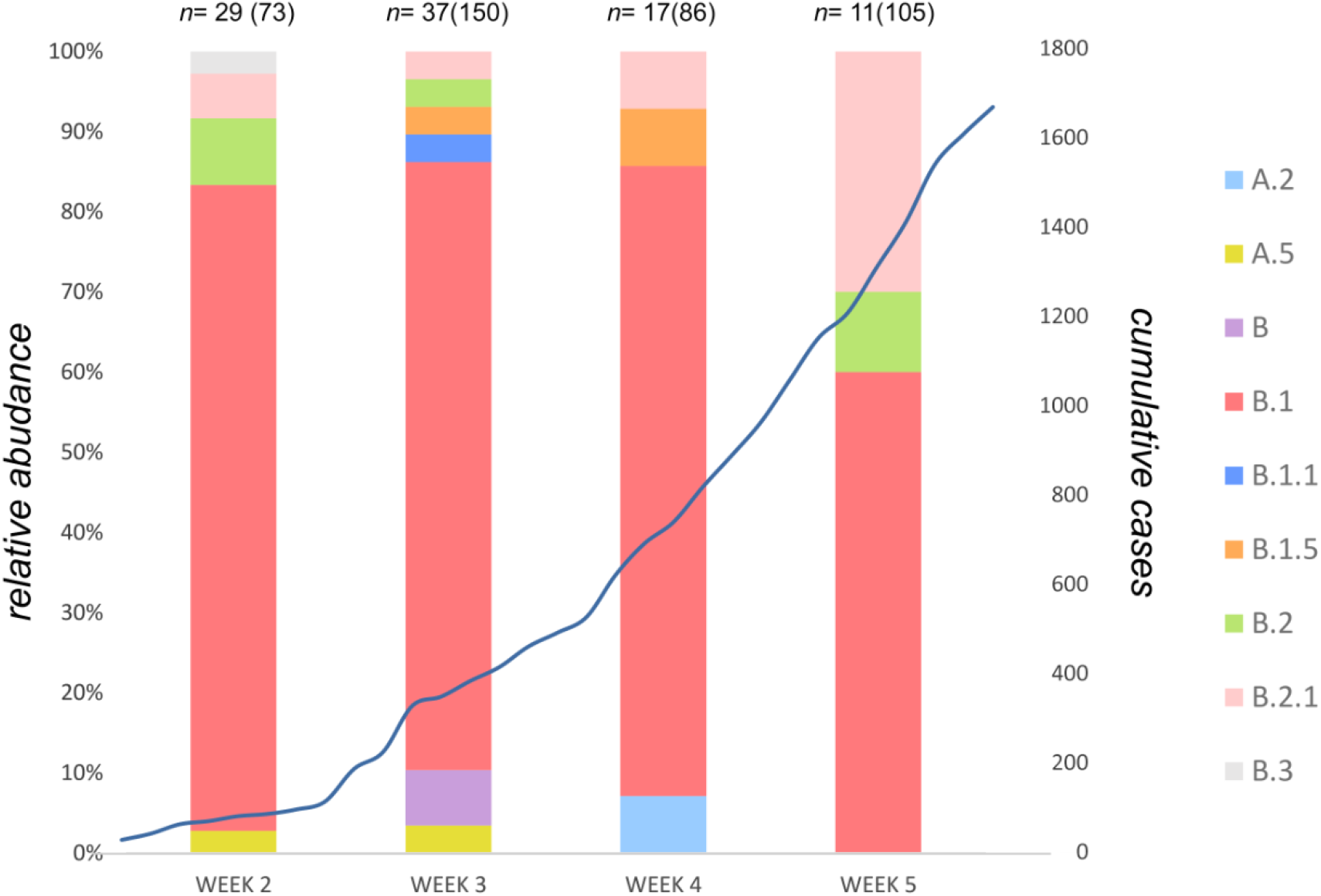
Stacked bar chart plotting the relative abundance of lineages between week 2 and week 5 of the outbreak in Athens, Greece. The overlaid linear graph represents cumulative confirmed COVID-19 cases in Greece between week 2 and week 5 of the outbreak. Numbers of cases per week that were included in this report are stated over the stacked bars. In the parenthesis are the total positive test per week reported by the Laboratory of Microbiology, Medical School, National and Kapodistrian University of Athens

**Suppl. Figure 3.**
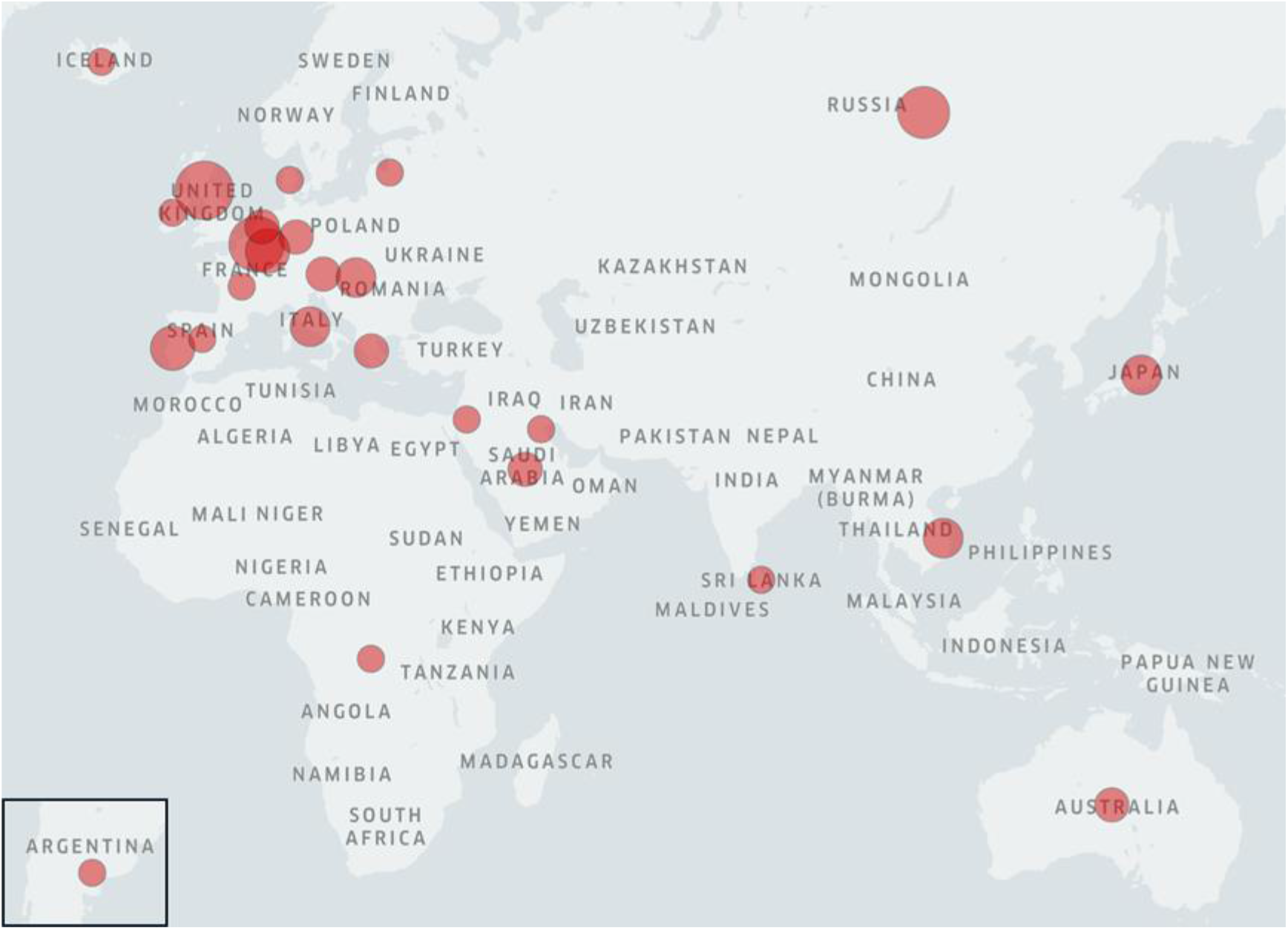
Map of the individual encounters of SARS-Cov2 variants without divergence from variants isolated in Athens, Greece. Spots represent the number of variants per country. Isolates (NextStrain) with lim x→0 pairwise distances with isolates included in this report were used for the illustration.

**Suppl. Figure 4.**
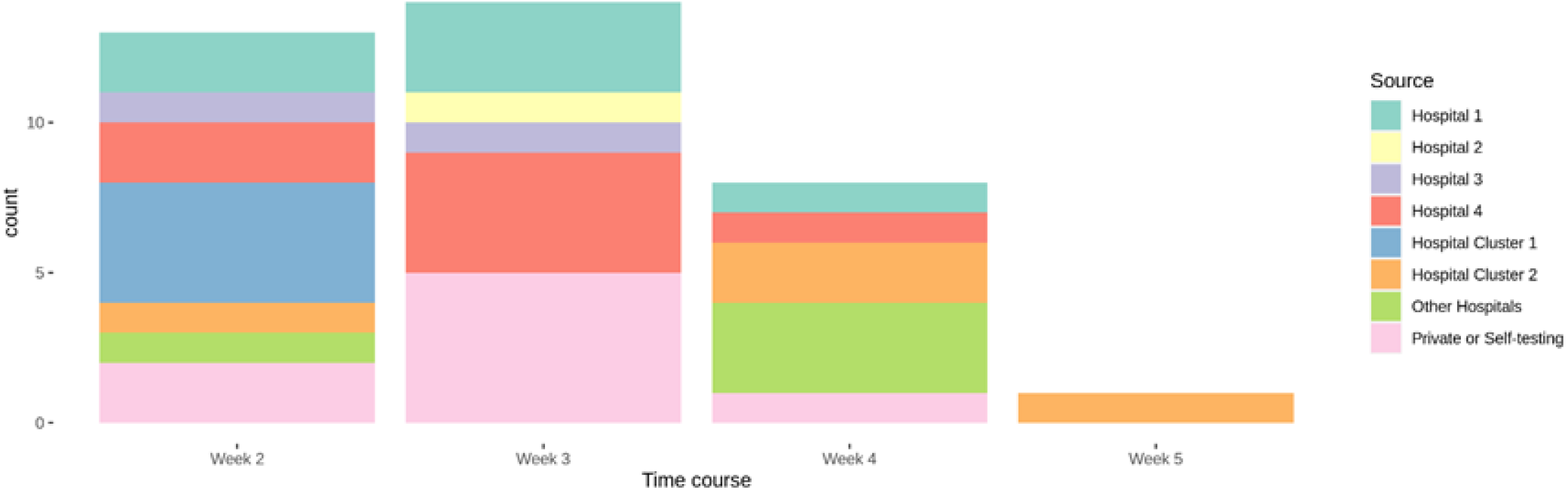
Stacked bar chart plotting the abundance of lineage B.1 dominant variant (n=36) between week 2 and week 5 of the outbreak in the health care providers of the study in Athens, Greece.

**Suppl. Figure 5.**
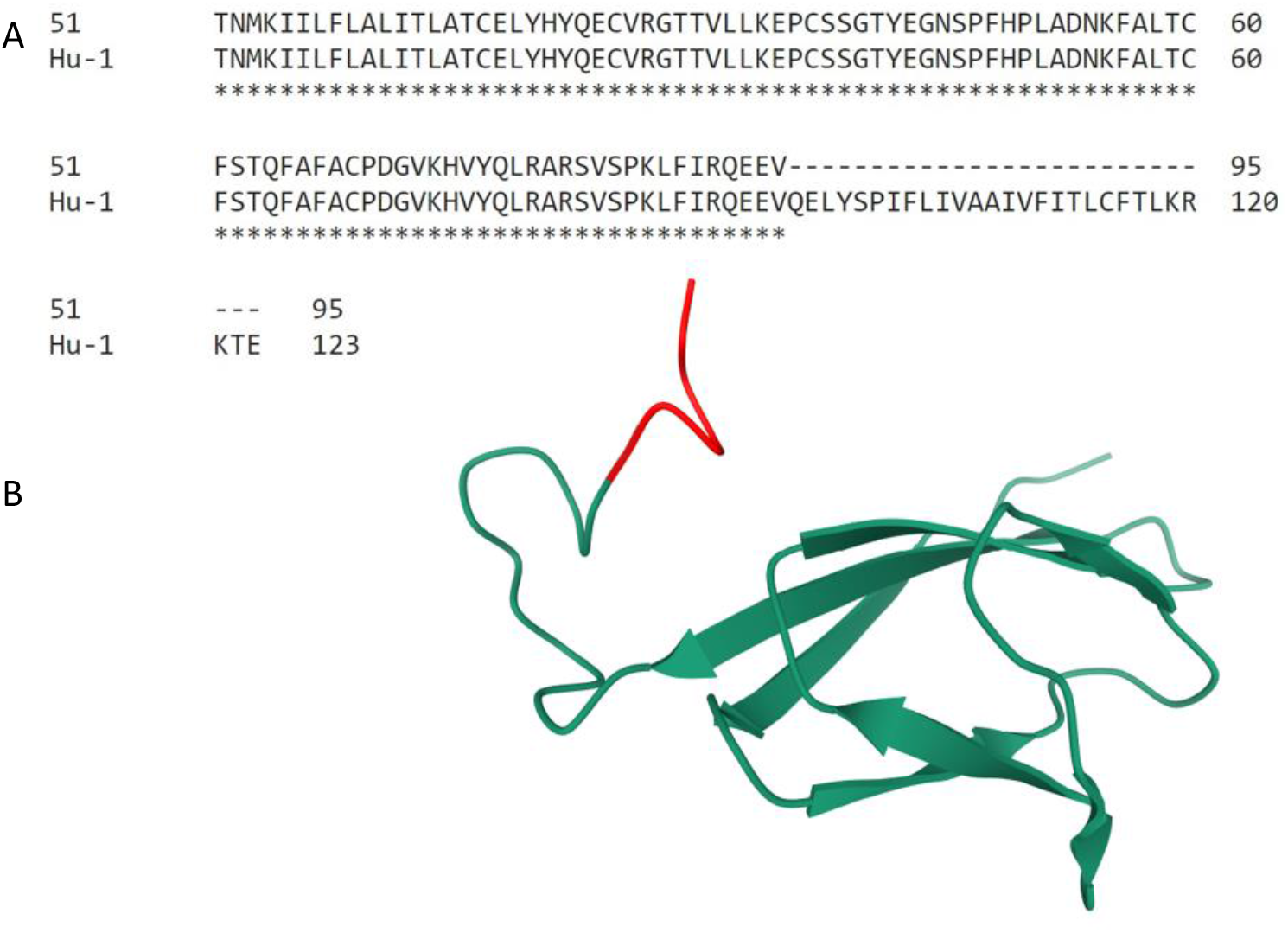
Isolate 51 premature stop codon resulted into abrogation of 28 amino acids from ORF7a protein. (A) Alignment of isolate 51 ORF7a amino acid sequence with reference SARS-Cov2 Hu-1 genome. (B) SARS coronavirus ORF7a protein 3D structure (Protein Data Bank ID: 1YO4) highlighting (red) the homologous amino acids predicted to be eliminated from isolate 51 SARS-Cov2 ORF7a protein.

## Supplementary Tables

**Supplementary Table 1.**
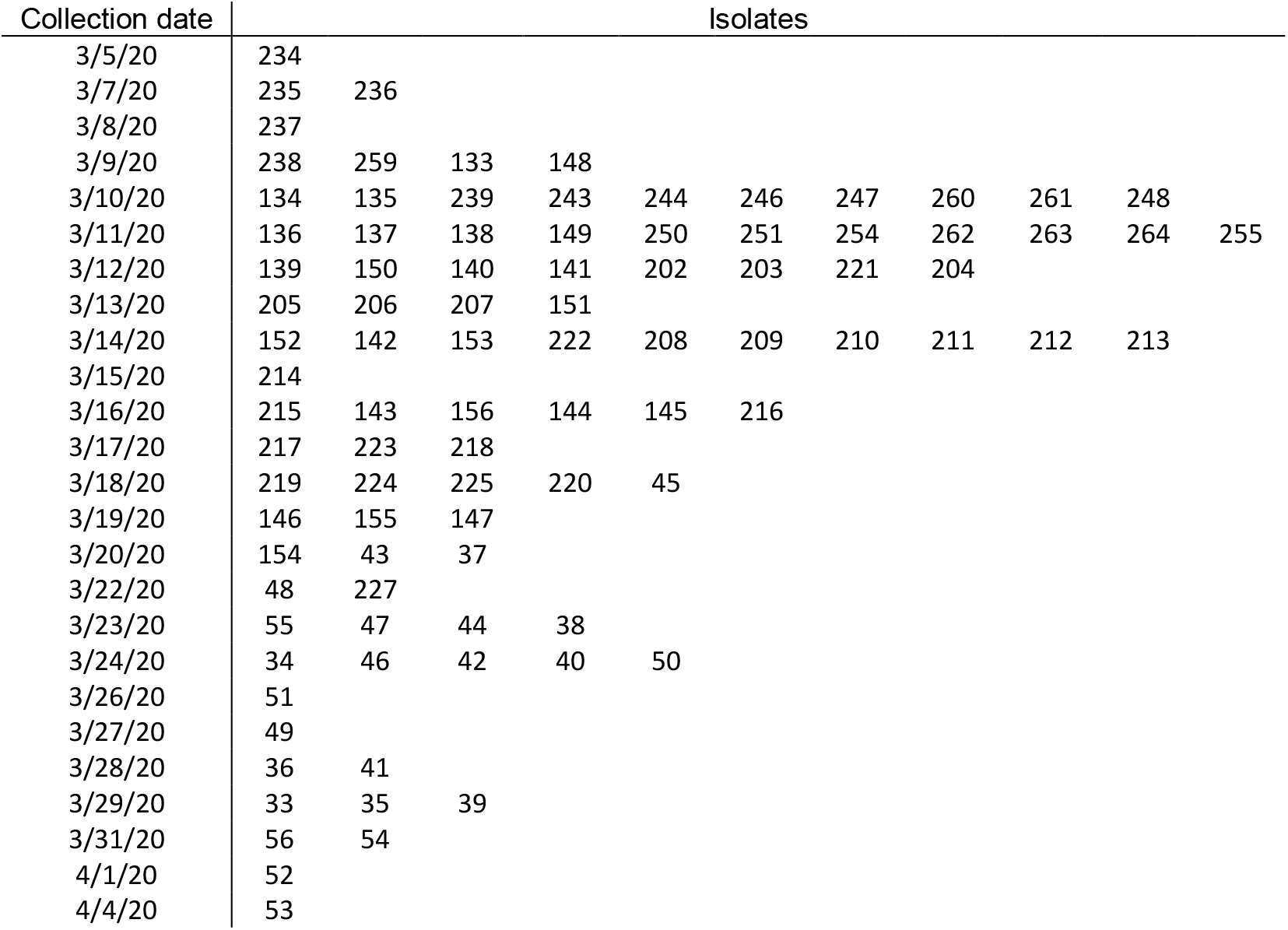
Collection dates of the 94 isolates analysed in the present study

**Supplementary Table 2.** Unique European and international isolates with minimum pairwise distances (<10^-12^) with isolates from Athens, Greece.

|  |  |
| --- | --- |
| Argentina/PAIS_A022/2020 | Kuwait/KU008/2020 |
| Australia/VIC1297/2020 | Latvia/011/2020 |
| Australia/VIC860/2020 | Luxembourg/LNS0945359/2020 |
| Austria/CeMM0098/2020 | Luxembourg/LNS3694003/2020 |
| Austria/CeMM0140/2020 | Luxembourg/LNS4836560/2020 |
| Belgium/HAA-030953/2020 | Luxembourg/LNS9470500/2020 |
| Belgium/ITM_C585/2020 | Netherlands/NA_138/2020 |
| Belgium/ULG-10013/2020 | Netherlands/NA_166/2020 |
| Belgium/ULG-10072/2020 | Portugal/PT0008/2020 |
| Belgium/ULG-10082/2020 | Portugal/PT0021/2020 |
| Belgium/ULG-6743/2020 | Portugal/PT0050/2020 |
| Belgium/ULG-7626/2020 | Portugal/PT0061/2020 |
| CzechRepublic/Seq6/2020 | Russia/Buryatia-87105/2020 |
| Denmark/ALAB-SSI286/2020 | Russia/Buryatia-88903/2020 |
| DRC/1398/2020 | Russia/StPetersburg-RII4328S/2020 |
| England/20100004806/2020 | Russia/StPetersburg-RII5169S/2020 |
| England/201360191/2020 | Russia/StPetersburg-RII5170S/2020 |
| England/BRIS-124C5C/2020 | Russia/StPetersburg-RII6060S/2020 |
| England/CAMB-76339/2020 | SaudiArabia/KAUST-Jeddah66/2020 |
| England/CAMB-7C54C/2020 | SaudiArabia/KAUST-Makkah163/2020 |
| France/ARA10251/2020 | Scotland/CVR305/2020 |
| Germany/BAV-MVP0101/2020 | Scotland/EDB2084/2020 |
| Germany/NRW-19/2020 | Scotland/EDB3846/2020 |
| Greece/135/2020 | Spain/Valencia218/2020 |
| Greece/155/2020 | SriLanka/COV53/2020 |
| Hungary/SRC-00105w/2020 | Taiwan/TSGH-03/2020 |
| Hungary/SRC-03670w/2020 | Vietnam/39607/2020 |
| Hungary/SRC-1788lc/2020 | Vietnam/OUCRU026/2020 |
| Iceland/467/2020 | Vietnam/OUCRU0419/2020 |
| Ireland/24052/2020 | Wales/PHWC-263ED/2020 |
| Italy/TE4880/2020 |  |
| Italy/TE5056/2020 |  |
| Italy/TE5551/2020 |  |
| Japan/P1/2020 |  |
| Japan/P2-2/2020 |  |
| Japan/P4-4/2020 |  |
| Jordan/SR-052/2020 |  |

